# Epidemic Models and Protective Mutations

**DOI:** 10.1101/2021.04.17.21254114

**Authors:** Claudio Parmeggiani

## Abstract

The paper proposes and discusses an epidemic model which assumes that a population, affected to a disease, is (partially) protected from the disease. These protections may be generated by some inherited genetic mutations; mutations favoured by the presence, in the past, of a deadly endemic disease (the Haldane hypothesis). The protections can also have a non-genetic origin: the cause can be the age or the sex of the subjects; in this case the model can only be applied for short periods, typically in the early stages of the epidemic. Anyway we shall see that the presence of these protections, if wisely managed, not only does it benefit the whole population, but it also makes the measures taken to contain the epidemic more effective.

## 1 Introduction

Here it is proposed and discussed an *epidemic model* which assumes that part of a population, affected to a disease, is, in some way and at different levels, *protected* from the disease. These protections may be generated by some inherited *genetic mutations*; mutations eventually favoured by the presence, in the past, of some deadly endemic disease (the Haldane hypothesis, see the Section 2). But these protections may very well have a *non-genetic* origin: the cause can be, say, the age or the sex of the subjects; in this case the epidemic model can, of course, be applied only for short periods, typically in the early stages of the epidemic. And you should take into account the circumstance that these protections can be rapidly lost, due to (random) counter-mutations of the disease agents.

Accordingly, in Sections 3 and 4, it is described a mathematical model which attempts to incorporate the above mentioned fact, that part of the affected population is, to varying degrees, protected; so, for example, while some individuals are fully susceptible, others are only mildly infected and weakly contagious. The Section 5 contains examples and numerical solutions of the differential equations of the model; you shall see that the protections, if wisely managed, not only does it benefit the whole population, but also makes the measures taken to contain the epidemic more effective. In the Appendices are outlined some mathematical techniques here used and are sketched some variant of the model.

The model can be regarded as a sort of extension of to the “classic” Kermack and McKendrick one [5] (see also [7]); here, as in [5], the disease is assumed to be spread only by *contact* and not, say, by the action of some vector. And, for certain matters, see also the still relevant Volterra “Lecons” [11]: the spread of the disease can be seen as a fight between an evolved DNA and a primitive RNA/DNA for the control of the “body”, body which both need to perpetuate themselves.

## 2 Protective Mutations

It is possible that at the onset of an epidemic, of a completely *new* type, a substantial part of the affected population is, partially or completely, protected? An answer (one of the most plausible, but not the only one, evidently) is that this part of the population has inherited some *protective mutations*, that is some *random* mutations, eventually slightly harmful, but strongly protective against an *old*, perhaps now eradicated, endemic disease. And also protective against the new one. This is one aspect of the so called “Haldane hypothesis” [3]:

> 1932 – The Causes of Evolution – What is Fitness? A study of the causes of death in man, animals and plants leaves no doubt that one of the principal characters possessing survival value is immunity to disease. Unfortunately, this is not a very permanent acquisition, because the agents of disease also evolve, and on the whole more rapidly than their victims. … It seems likely that when a species is subjected to a series of attacks by an evolving parasite it may be forced along a path of structural change by its temporarily successful acquisition of immunity. But in the end it may driven, so to say, into a corner, where further immunity involves structural changes which are disastrous to it in its everyday life.

More explicitly, always Haldane, in [4]:

> 1949 – Disease and Evolution. I want to suggest that the struggle against disease, and particularly infectious diseases, has been a very important evolutionary agent, and that some of its results have been rather unlike those of the struggle against natural forces, hunger, and predators, or with members of the same species. Under the heading infectious disease I shall include, when considering animals, all attacks by smaller organism, including bacteria, viruses, fungi, protozoa, and metazoan parasites.

Evidently the old disease, to have given rise to effective mutations, must have been endemic, long lasting (hundreds of generations) and, if now eradicated, its disappearance must not have been too far in time. For the human species, on Earth, a truly plausible candidate for this role is *malaria* (“aria mala”, evil air): for thousands of years it has spread throughout the planet and has marked the history of peoples. And even when, after a long struggle, it was eradicated, all this occurred in recent times, in the last century; consider, for example, the case of Italy [10] or of Taiwan [6].

Unfortunately the informations about the *genetic legacy* of malaria are very far to be systematized and complete; but see, for example, [12] (WHO 2019) or Carter & Mendis [2]. In this regard it is interesting to note that, apparently, the genetic legacy linked to the presence of *P. falciparum* (predominant in Sub-Saharan Africa) is quite different from the one related to the presence of *P. vivax* (mainly widespread in East Asia, Europe and Middle East, in the past).

## 3 The rules

### 3.1 Infectible, infected, recovered or deceased

Let *N* (t) be the size of some population 𝒩 (a set of *individuals*) living, at time *t*, in a fixed region. Part of the population is affected by some *new* contagious disease and can spread it; taking into account the fact that the disease affects individuals in different ways, the set 𝒩 is partitioned into several disjointed classes, (𝒩_*k*_)_*k* ∈*K*_; the members of each class are assumed to react differently to infection. *K* is here a *finite set* (but see the Appendix A); |*K*| will denote the number of elements of *K*.

At the onset of the epidemic each 𝒩_*k*_ can be split into *two* subclasses, the *uninfected* 𝒳_*k*_, which are, in varying degrees, capable to be infected; and the *infected*, 𝒴_*k*_, which can eventually transmit the disease. Then, say, different 𝒳_*k*_ have different degrees of *susceptibility*: at one extreme there are totally susceptible individuals, at the other extreme individuals who can never catch the disease, they are fully *protected*. Correspondingly, members of different 𝒴_*k*_ present different severity of the infection and variable *infectiousness* and *evidence of symptoms*. We shall denote by 𝒳 (𝒴) the *union* of all the 𝒳_*k*_ (𝒴_*k*_); 𝒩 is the union of 𝒳 and 𝒴.

Besides, during the epidemic, some individual can *acquire*, after recovering, an antibody driven (typically temporary) *immunity*; these immunities can sometime accumulate in complicate ways, but here we are only marginally interested to them; we shall simply introduce a new subset of 𝒩, *Ƶ* (the *immune*) and its subclasses, (*Ƶ*_*k*_)_*k* ∈ *K*_, whose members are recovered individuals, from 𝒴_*k*_, now assumed to be *totally* but *temporarily* immune. The *Ƶ*_*k*_ are uninfected, as the 𝒳_*k*_, which are also non-immune and can now be more properly referred to as *infectible*. These immunities can also be *induced*, by external intervention; induced immunities are, generally, longer lasting; those ones will not considered here.

If *n*_*k*_ (*t*), *x*_*k*_ (*t*), *y*_*k*_ (*t*) and *z*_*k*_ (*t*) are the sizes (number of individuals) of 𝒩_*k*_, 𝒳_*k*_, 𝒴_*k*_ and *Ƶ*_*k*_; and if *X* (*t*), *Y* (*t*) and *Z* (*t*) are the sizes of 𝒳, 𝒴 and *Ƶ* (everything at time *t*):

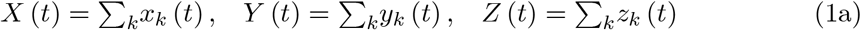

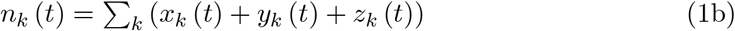

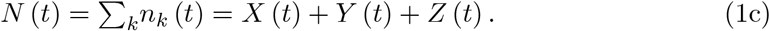

### 3.2 The spread of the disease

The decrease of the number of the uninfected, *x*_*h*_, *h* ∈ *K* (and the corresponding increase of the number of the infected, *y*_*h*_), per unit of time, due to *infectious contacts*, is assumed to be proportional to *x*_*h*_; that is to be equal to *c*_*h*_ (*t*) *x*_*h*_ (*t*) where *c*_*h*_ (*t*) is the (infection) *spread rate* – always ≥ 0. *c*_*h*_ generally depends on the likelihood of meeting contagious individuals; if only the infected (not the recovered) can spread the disease, we can assert that, in a first, linear approximation (*r*_*h,k*_ (*t*) ≥ 0, *h, k* ∈ *K*),

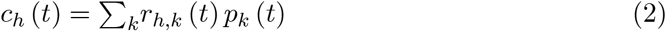

where the *p*_*k*_ (*t*) are the *probabilities* that a 𝒳_*h*_ (an uninfected member of 𝒳_*h*_) meets an (infected and infectious) 𝒴_*k*_.

Assuming that the distributions of infected is quite uniform – but this is not always the case – the *p*_*k*_ (*t*) are simply equal to *ŷ*_*k*_ (*t*) = *y*_*k*_ (*t*) /*N* (*t*), the *density of infected*; 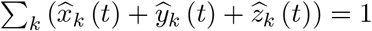. Thus we can conclude that the decrease of *x*_*h*_ (*increase* of *y*_*h*_), per unit of time (per day, per week …), due to infectious contacts, is equal to:

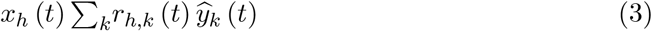

and that the analogous decrease of *X* (increase of *Y*) is:

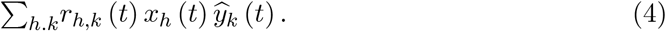

If | *K* | = 1 the above formulae reduce to the well know one, *r* (*t*) *X* (*t*) *Y* (*t*) /*N* (*t*).

The *contact matrix* (*r*_*h,k*_)_*h,k*∈*K*_ may significantly vary over time due to *external* interventions aimed, for example, at preventing uninfected-infected contacts; in most case *r*_*h,k*_ (*t*) can be simply factored as *r* (*t*) *γ*_*h,k*_ (or as *r*_*h*_ (*t*) *γ*_*h,k*_) where the *γ*_*h,k*_ depend only on the disease characteristics and are *constant*, for a stable disease agent. The *contact rate r* (*t*) is here normalized by setting the *maximum* of the *γ*_*h,k*_ equal to 1.

Not all the infected can contact others individuals: some of them are *isolated*. This fact can be handled introducing, for every *k* ∈ *K*, the *isolation weights, q*_*k*_ (*t*), ≥ 0 and ≤1; *q*_*k*_ (*t*) = 1 would mean that all the individuals of 𝒴_*k*_ are isolated; we shall always assume that all the *q*_*k*_ (*t*) are < 1; ∑_*k*_ *q*_*k*_ (*t*) *y*_*k*_ (*t*) < *Y* (*t*). Therefore, in the above formulae, *ŷ*_*k*_ (*t*) must be replaced with:

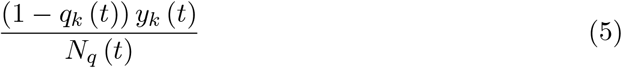

where *N*_*q*_ (*t*) := *N* (*t*) −∑_*k*_ *q*_*k*_ (*t*) *y*_*k*_ (*t*).

Even the *q*_*k*_ may vary over time due to *external* interventions, aimed, in this case, to detect the presence of *asymptomatic* individuals. But sometimes it is not so easy to recognize the presence of the infection, it is then necessary to do some time (and resources) consuming tests and to organize and regulate the screening activities, all not very popular things. So these activities are often carried out when it is too late, when there are too many infected.

Moreover the number of infected decrease because some of them will recover or die (or they are suppressed or extirpated, being “expendable”); hence, if *a*_*k*_ (*t*) and *d*_*k*_ (*t*) are, respectively, the *recovery* and the (infection related) *deaths rates* (dead-plus-suppressed),

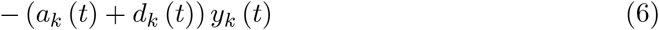

is the corresponding change of *y*_*k*_ (*t*), per unit of time. Then, if *b*_*k*_ (*t*) is the rate of (acquired) *immunity loss*, the change of *z*_*k*_ (*t*), per unit of time, is:

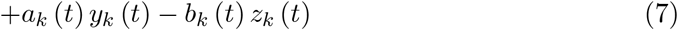

while the consequent changes of *x*_*k*_ (*t*) is:

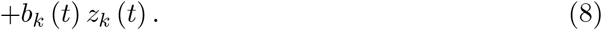

Generally *a*_*k*_ + *d*_*k*_ > 0, but *a*_*k*_ = *d*_*k*_ = 0 for *disease tolerant* individuals: infectible individuals, eventually infectious when infected, but always “healthy”: they *coexist* with the disease agents.

The *a*_*k*_ and *d*_*k*_ may vary over time, sometimes dramatically, due to external causes (improvements in therapies or, for example, collapse of healthcare structures); the *b*_*k*_, on the contrary, are generally more constant.

If *a*_*k*_ + *d*_*k*_ is > 0, the *τ*_k_ = 1/ (*a*_*k*_ + *d*_*k*_) can be understood as the *average durations* of the infection: in that period of time an average infected (𝒴_*k*_) heals or dies. The dimensionless “indices” *τ*_*k*_*a*_*k*_ and *τ*_*k*_*d*_*k*_ are, respectively, the *probabilities* that someone, after contracting the disease, recover or die, by the disease; *τ*_*k*_*a*_*k*_ + *τ*_*k*_*d*_*k*_ = 1, how it should be. Likewise the *σ*_*k*_ = 1/*b*_*k*_ (if *b*_*k*_ is > 0) are the average durations of the acquired immunity; if *σ*_*k*_ is of the order of magnitude of the average life span of the population, the immunity can be regarded as permanent.

Note that if |*K*| is > 1 the increase of *y*_*k*_ is *not proportional* to *y*_*k*_ and the dimensionless parameters *rτ*_*k*_ (or *r*_*k*_*τ*_*k*_) lose, as they are, their meaning. See the Appendix B, The infected.

Finally, for long-lasting epidemics (when the protections are of genetic origin), we have to consider the changes of number of individuals due to *births* and *natural deaths*. It can happen that the newborns are infected or immune; assuming that they are not, we can say that these changes − births, *ν* and deaths, *μ* − are, per unit of time and for each one of the subclasses 𝒳_*k*_, 𝒴_*k*_ and *Ƶ*_*k*_, simply equal to:

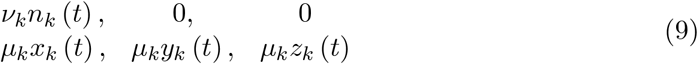

The deaths rates are here assumed to be the same for the 𝒳, 𝒴 and *Ƶ*; *ν*_*k*_ and *μ*_*k*_ are constant, *ν*_*k*_ ≥ 0, *μ*_*k*_ > 0; it is generally not necessary to introduce a carrying capacity for the *ν*. In some case the time period examined is relatively short − if, for example, we are only interested to the initial stages of the epidemic − and the *ν* and *μ* can be ignored.

## 4 The equations

Here *I* is the real interval [0, +∞ [; the *linear space E* = ℝ^*K*^ is normed by, say, ‖*u*‖ =∑_*k*_ |*u*_*k*_|;

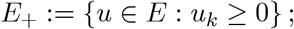

*a, b, d, q* are *continuos* and *bounded* maps *I* → *E*_*+*_, *q*_*k*_ < 1; the *r*_*h,k*_ are continuos and bounded functions *I* → ℝ_+_, Σ_*h,k*_ *r*_*h,k*_ > 0.

Translating the “changes per unit of time” with derivatives with respect to *t*, we arrive at a system of *ordinary differential equations*, in ***x, y*** and ***z*** (***n** = **x** + **y** + z*):

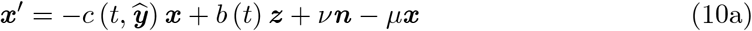

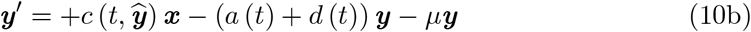

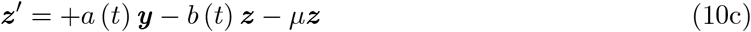

where ***x, y*** and ***z*** are *continuously differentiable* maps *I* → *E*_+_ assuming some fixed value when *t* = 0. Moreover (*λ*_*k*_ := *ν*_*k*_ − *µ*_*k*_):

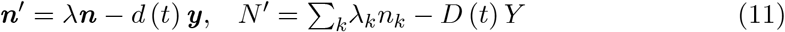

and

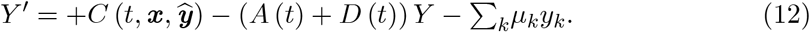

Above, if *a* and ***x*** ∈ *E, ax* stands for (*a*_*k*_*x*_*k*_)_*k*∈*K*_, ***ŷ***= (***y*** − *q* (*t*) ***y***) /*N*_*q*_ (*t*),

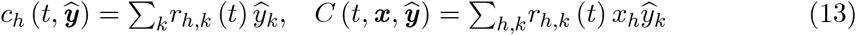

and *A, D* are defined by *A* (*t*) *Y* = Σ_*k*_*a*_*k*_ (*t*) *y*_*k*_, *D* (*t*) *Y* = Σ_*k*_*d*_*k*_ (*t*) *y*_*k*_.

We can eventually assume that the maps *a, b, d, q* and *r* are only *measurable* and *bounded* and that the functions ***x, y*** and ***z*** are only *absolutely continuos* – hence almost everywhere differentiable. For this and all the other mathematical aspects of the problem, see Birkhoff & Rota [1] and Sansone & Conti [8].

When *t* = 0 (the *epidemic onset*) *all* the *n*_*k*_ and *x*_*k*_ are assumed to be > 0 (typically *x*_*k*_ (0) ≅ *n*_*k*_ (0)); it follows that all the *n*_*k*_ (*t*) and *x*_*k*_ (*t*) are *always* > 0, for t ≥ 0. See the Appendix B for this and the following remarks.

*Y* (0) also has to be > 0 (otherwise *Y* (*t*) would be identically = 0), *Y* (*t*) is then always > 0. But, for some *k* ∈ *K, y*_*k*_ (0) can be = 0; generally, in this case, *y*_*k*_ (*t*) > 0, for *t* > 0. However it may be that instead *y*_*k*_ (*t*) remains = 0 (so *x*_*k*_ (*t*) = *n*_*k*_ (*t*)): these individuals are not really taking part in the epidemic, they are fully protected.

*Z* (0) is obviously = 0, but, apart from special cases (*a*_*k*_ = 0 or *y*_*k*_ = 0), *z*_*k*_ (*t*) is > 0, for *t* > 0.

Note that the (***x, y, z***) differential system is very far from being *autonomous:* the *r* and the *q* are frequently varied along the epidemic in a planned and controlled way, having decided what to maximize and what to minimize. However, it can happen – mainly at the epidemic onset – that *r* and *q* are instead capriciously modified, following the results of opinion polls or the decrees of some autocrat.

In any case, in Section 5, will be examined some simple case; the equations are *numerically integrate* choosing two different, somehow opposite, planning and control strategies for the contact rate and the isolation weights. It will also be examined the case of *deadly diseases* (*a*_*k*_ = 0 and *d*_*k*_ > 0 for some *k* ∈ *K*) and the *endemic phase* of the disease – when the density of infected, *Y/ N*, is, so to speak, *sensibly constant*.

## 5 Numerical integrations

In all examples of Sections 5.1 and 5.2, *K* = {*S, P*}: the *x*_*S*_ are totally susceptible, unprotected individuals; the *x*_*P*_ are less susceptible and partially protected (they cannot dye by the disease); *r*_*h,k*_ (*t*) = *r* (*t*) *γ*_*h,k*_;

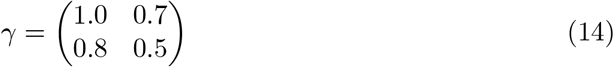

and (D is some unit of measure of time, say *days*)

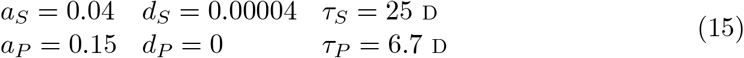

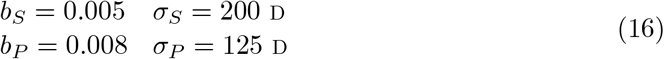

The probability that an unprotected individual dies, after contracting the disease, is equal to *d*_*S*_*τ*_*S*_ = 0.001. The *a, b* and *d* are assumed to be constant; the contact rate *r* and the isolation weights *q*_*S*_ and *q*_*P*_ are instead *variable*:

– when 0 ≤ ***t*** < ***t***_1_ the presence of the disease is *not perceived* (or, for “some reason”, underestimated) and:

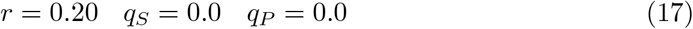
– when ***t***_1_ ≤ ***t*** < ***t***_2_ two different strategies are adopted. In the first one *r* is slightly decreased and *kept constant*; the tracking and screening activities are *enhanced*, so that, say:

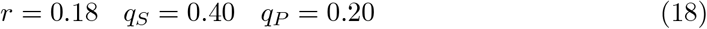 In the second strategy *r* is *continuously adapted* to *Y/N* (via the *function F*) while the isolation weights are less markedly increased:

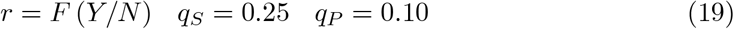
– when ***t***_2_ ≤ ***t*** ≤ ***t***_*M*_ it is assumed, *hopefully* but perhaps unwisely and surely recklessly, that the epidemic is about to end. Consequently the pre-disease value of *r* is restored while *q*_*S*_ and *q*_*P*_ are kept at quite low levels:

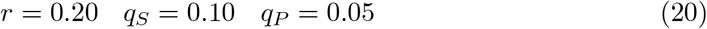

### 5.1 Few protected individuals

Now we have to fix the *initial conditions*; it is here assumed that only *one third* of the population is protected; and that, when *t* = 0, there are 10^−6^*N*_0_ (*N*_0_ := *N* (0)) infected individuals (protected):

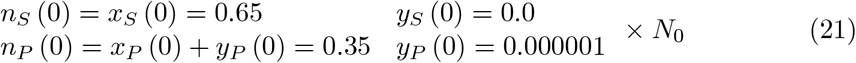

As always in this Section, *ν*_*S/P*_ = 0, *µ*_*S/P*_ = 0 and *z*_*S/P*_ (0) = 0.

Keeping *r* constant (= 0.18) for 90 D ≤ t ≤ 900 D (the *first strategy*), we arrive at the graph of **Fig. 1a**: after the initial peak, the infected curve flattens out, through *some damped oscillations* (sometimes improperly called “waves”.)

**Fig. 1a.**
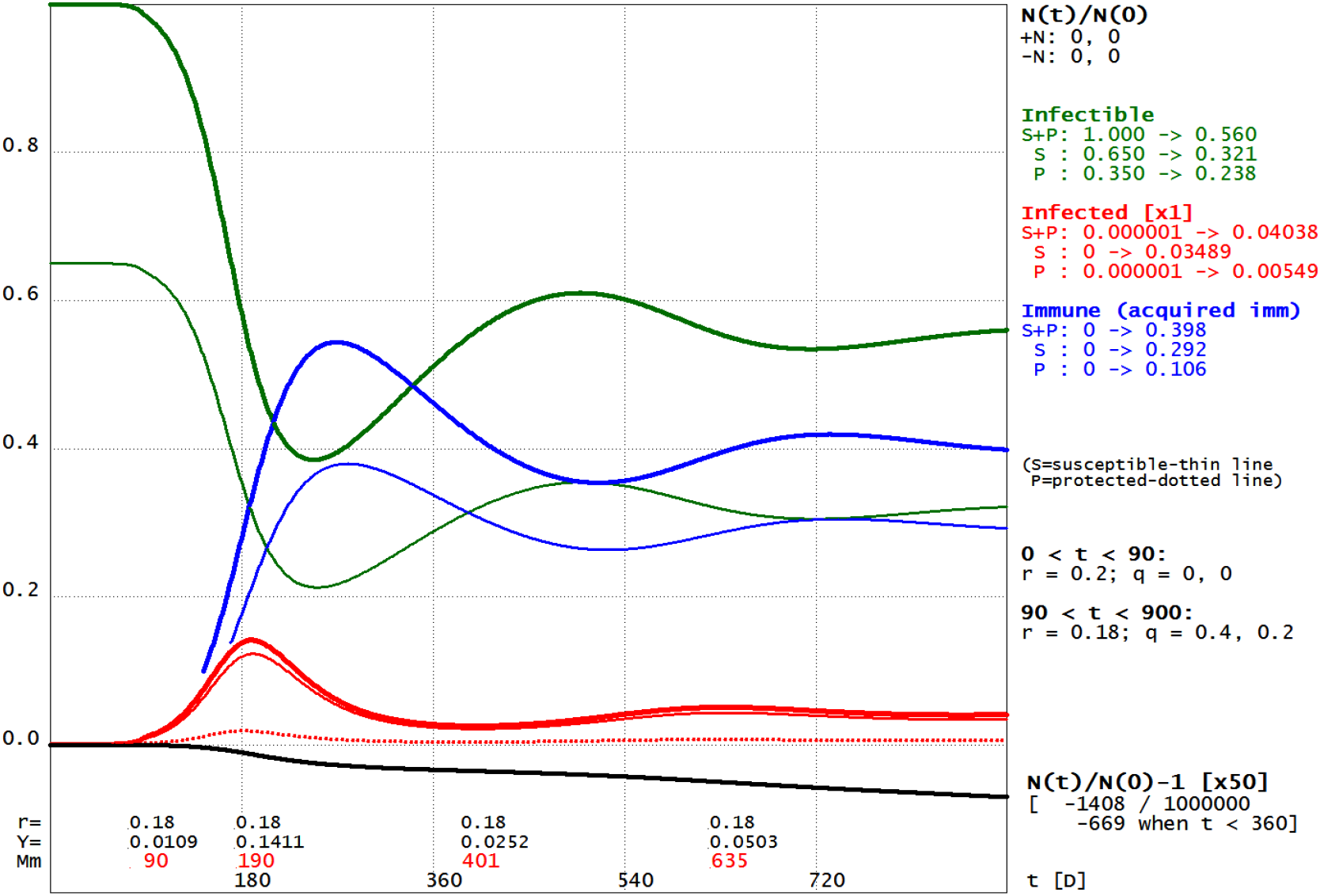
*x*_*S*_ (0) = 0.65, *x*_*P*_ (0) = 0.35. *r* is decreased from 0.20 to 0.18, *q* = 0.4, 0.2.

But if, following the *second strategy, r* is continuously adapted to *Y*/*N* – the total number of infected – by, for example,

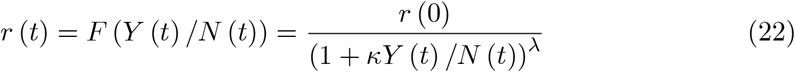

with *κ* = 3 and *λ* = 2 (always for 90 D ≤ t ≤ 900 D) you will get the graph of **Fig. 1b**.

**Fig. 1b.**
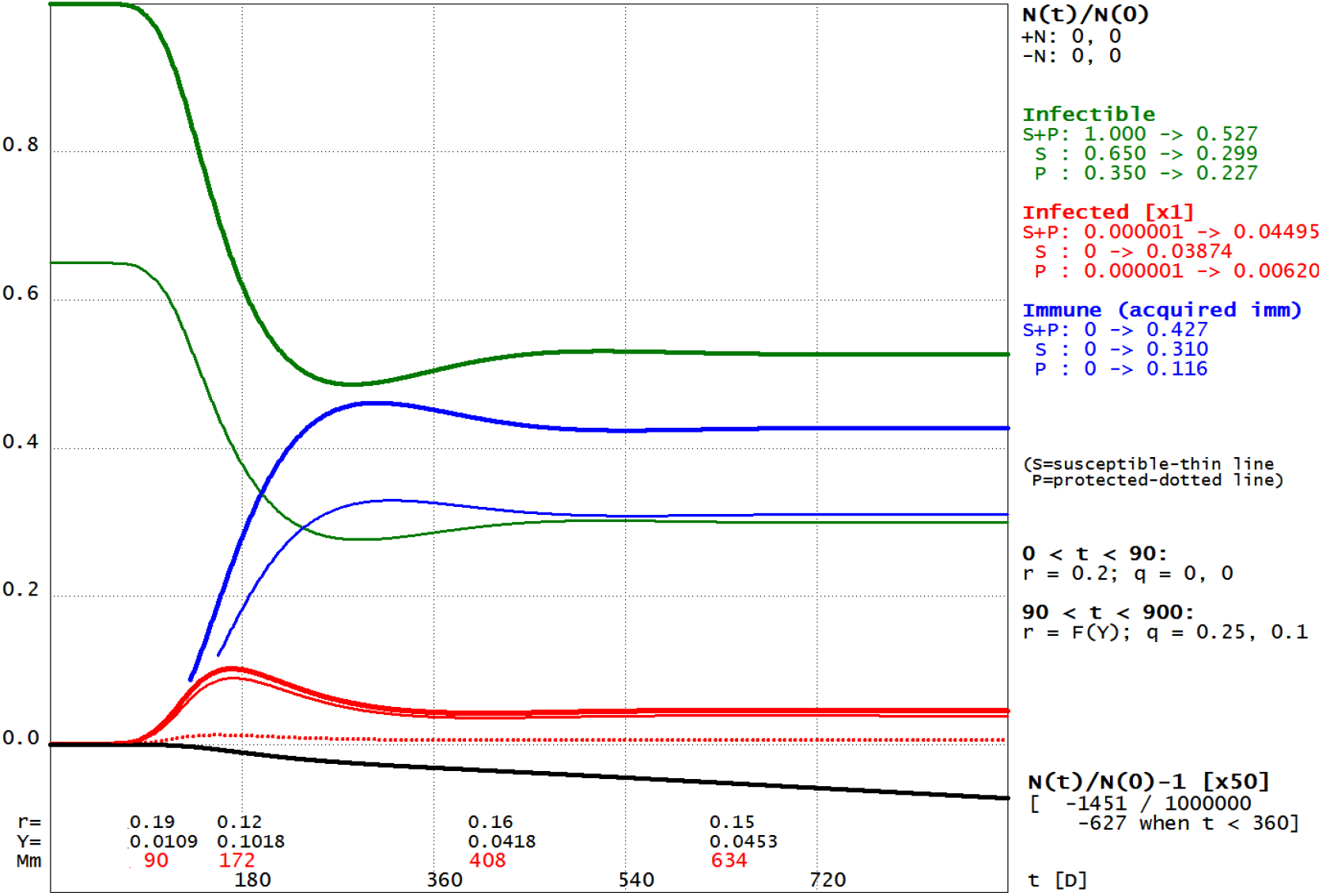
*x*_*S*_ (0) = 0.65, *x*_*P*_ (0) = 0.35. *r* (0) = 0.20, *r* = *r* (0) / (1 + 3*Y*)^2^, *q* = 0.25, 0.1

You can see, as it was easily predictable, that the infected curve is now more smooth and flat (at the curve peak, *r* = 0.12), but the “results” are, more or less, the same of **Fig. 1a**. Obviously a flatter curve places less stress on healthcare facilities; but the continuous adaptation, more or less forcibly, of *r* to *Y*/*N* has a considerable social and economic cost.

What happens if, from a certain point in time (*t* ≥ 345 D), the pre-disease parameters are restored? You will inevitably return to the starting point (*t* = 0, “back to square one”), now clearly with more infected and (fortunately) with more immune too; moreover this second, “artificial” peak, is quite similar to the first.

See **Fig. 1c**, where it is taken into consideration the second strategy; for the first one the situation is similar, but slightly better: there are *more immune* individuals.

**Fig. 1c.**
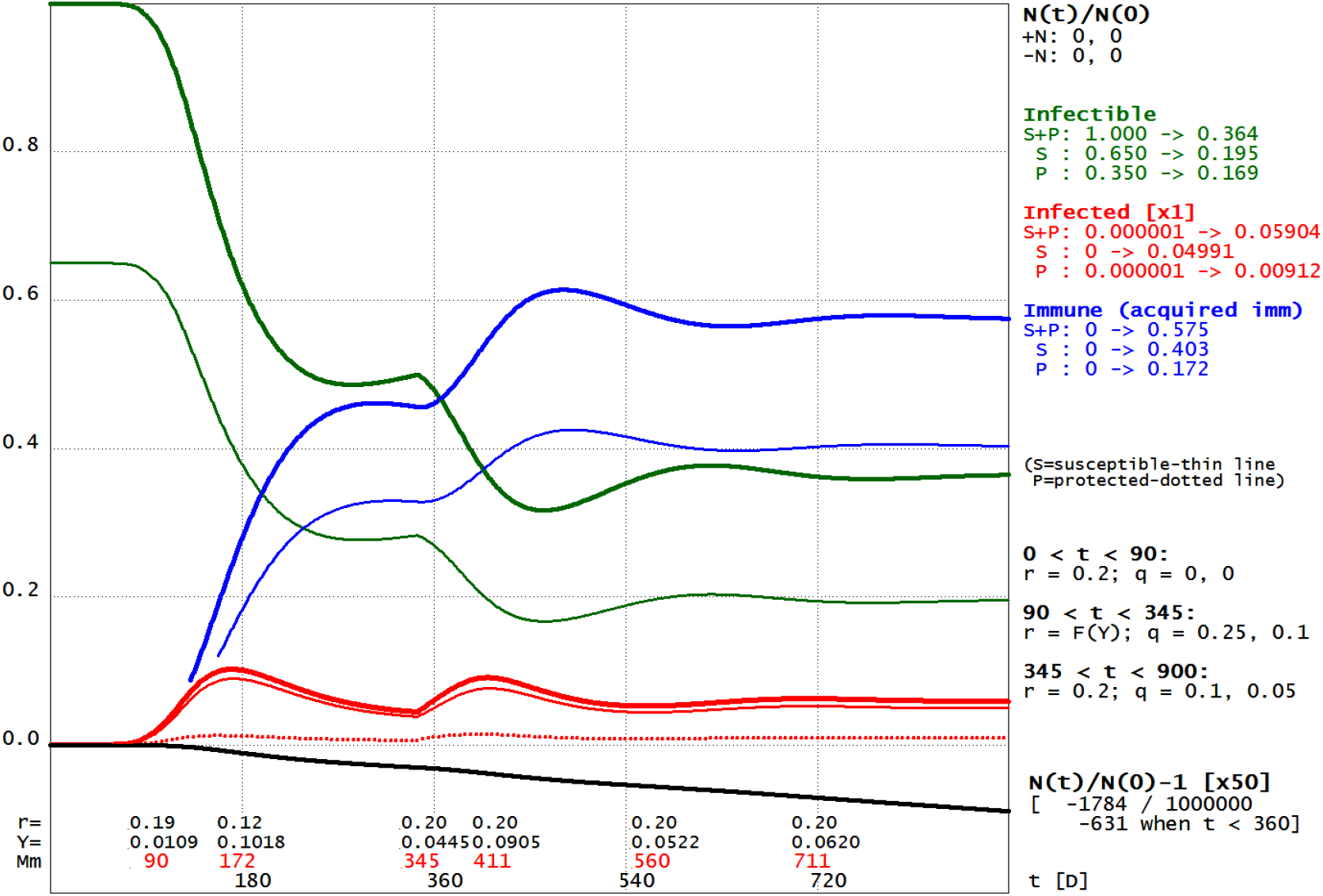
*x*_*S*_ (0) = 0.65, *x*_*P*_ (0) = 0.35; *r* = 0.20/ (1 + 3*Y*/*N*)^2^. For t ≥ 345, *r* = 0.20.

### 5.2 Many protected individuals

Now *two thirds* of the population are protected and, when *t* = 0, there are 2 × 10^−6^*N*_0_ infected individuals (protected):

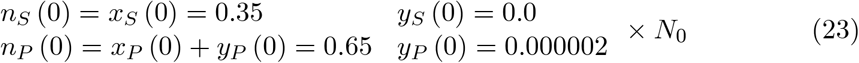

Again *ν*_*S/P*_ = 0, *µ*_*S/P*_ = 0 and *z*_*S/P*_ (0) = 0.

The situation is obviously more favorable; we can now intervene in a very soft way, for example keeping *r* = 0.20 and setting *q*_*S*_ = 0.25, *q*_*P*_ = 0.10 (when *t* ≥ 90 D) without serious consequences; see **Fig. 2a**.

**Fig. 2a.**
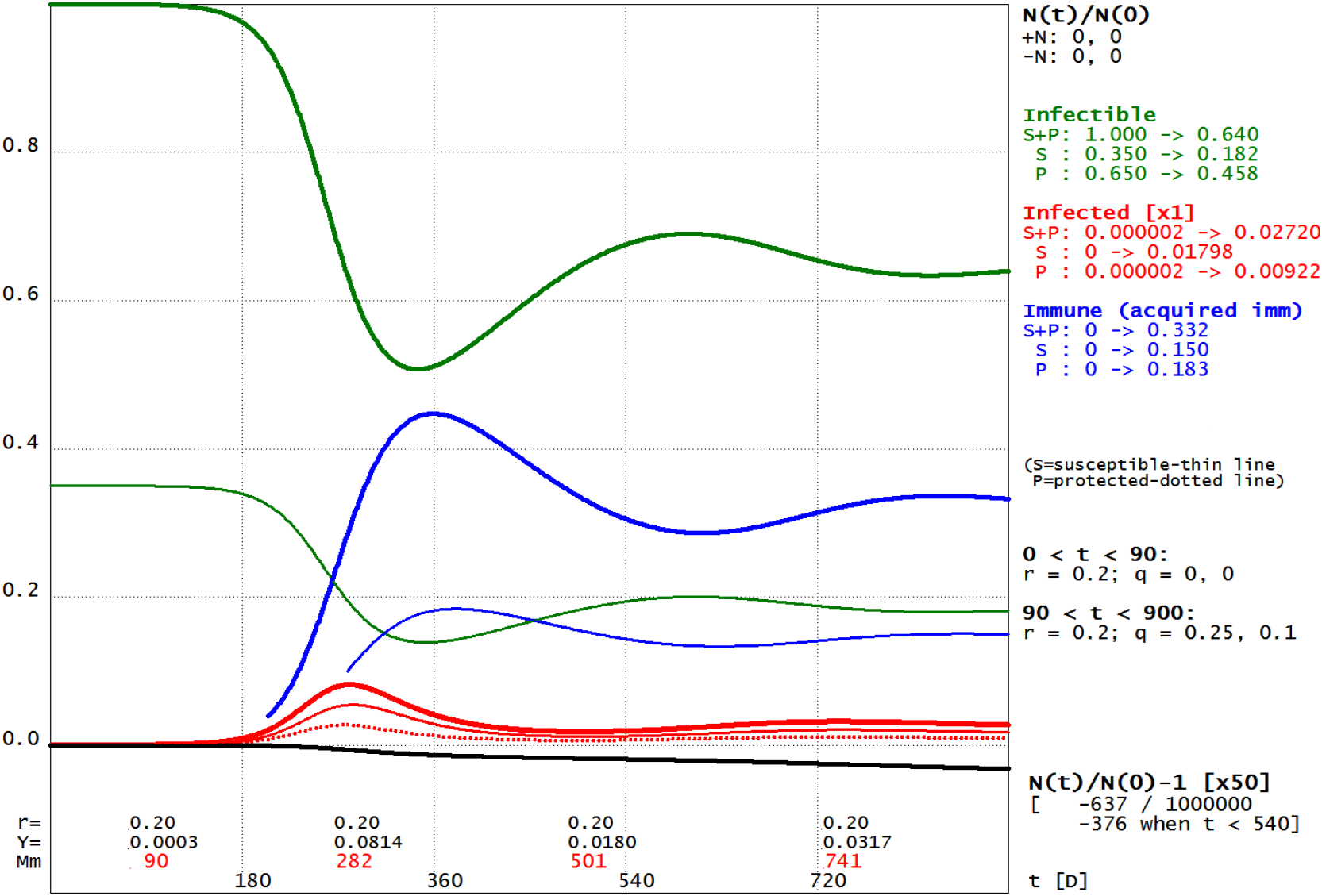
*x*_*S*_ (0) = 0.35, *x*_*P*_ (0) = 0.65. *r* is kept equal to 0.20, *q* = 0.25, 0.1.

A decidedly more energetic intervention, *r* = 0.18, *q*_*S*_ = 0.40 and *q*_*P*_ = 0.20 (always when *t* ≥ 90 D) can in fact almost suppress the infection; see **Fig. 2b**. But there is a catch: when the initial conditions are restored, the number of immune is still small and the infected and immune curves will have a marked peak, as in **Fig. 2c**.

**Fig. 2b.**
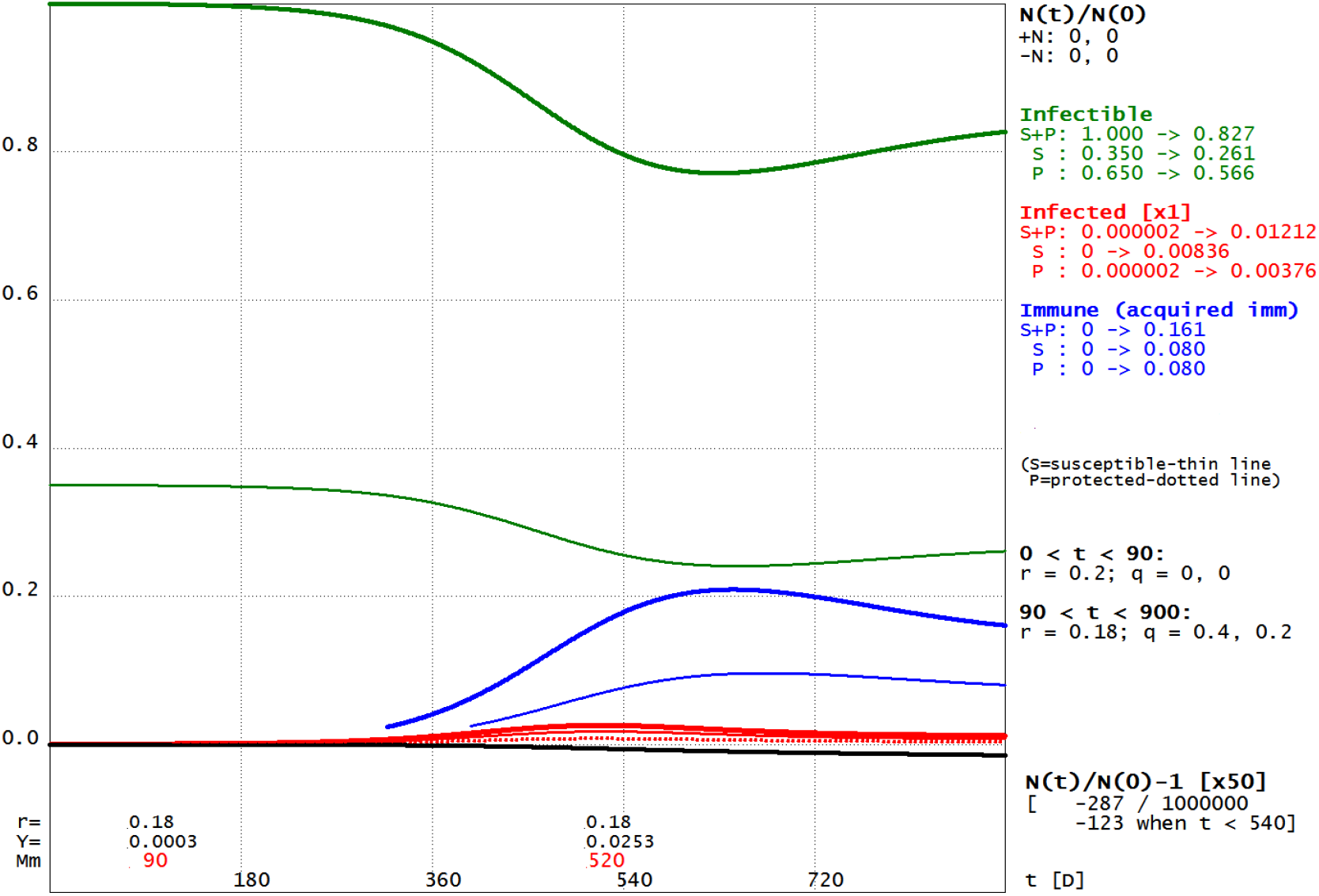
*x*_*S*_ (0) = 0.35, *x*_*P*_ (0) = 0.65. For *t* ≥ 90, *r* = 0.18, *q* = 0.4, 0.2.

**Fig. 2c.**
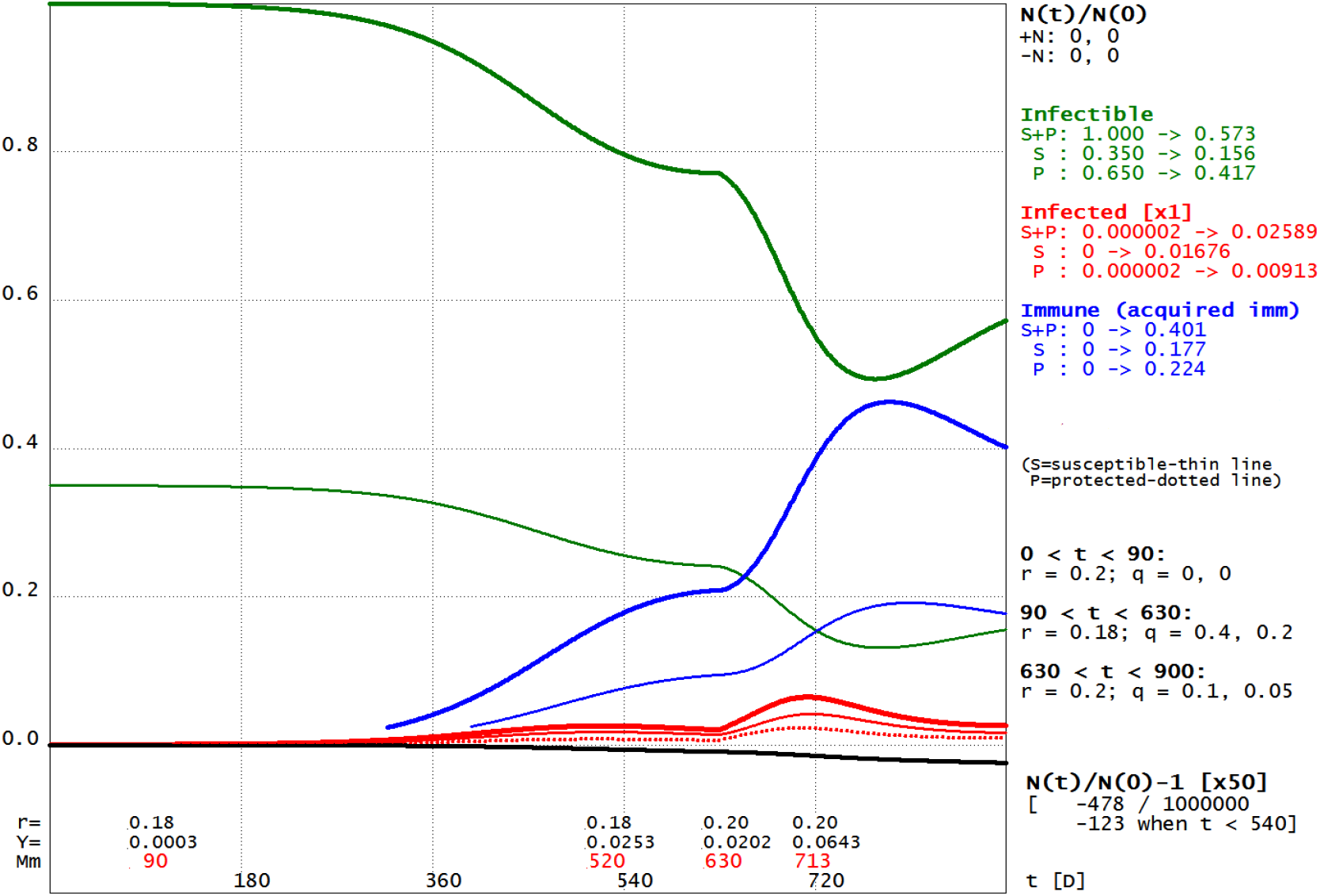
*x*_*S*_ (0) = 0.35, *x*_*P*_ (0) = 0.65. *r* (= 0.20) will be restored when *t* = 630.

Observe that the initial peak can be, by a *very* energetic intervention, *completely eliminated*. If, in the example of **Fig. 2b** we set, when *t* = *t*_1_ = 90 D, *r* = 0.14, then *Y* (*t*) ≤ *Y* (*t*_1_) = 0.00029*N*_0_ (for *t* ≥ *t*_1_) and the deaths number due to the disease is almost nil. But sooner or later you will have to reset *r* to its pre-disease value and thus generate the usual peak of infected – unless, in the meantime, a way is found to “produce” a substantial number of *immune* individuals.

### 5.3 Deadly Diseases

If, for some *k* ∈ *K, a*_*k*_ = 0 and *d*_*k*_ > 0 (a *deadly disease*, for the *x*_*k*_), then *z*_*k*_ = 0 (Appendix B); moreover, if *ν*_*k*_ and *µ*_*k*_ are ignored, *x*_*k*_ *monotonically decreases*. But now we are interested to a more realistic case so, in the following example (**Fig. 3a**), *K* = {*S, P*} and

**Fig. 3a.**
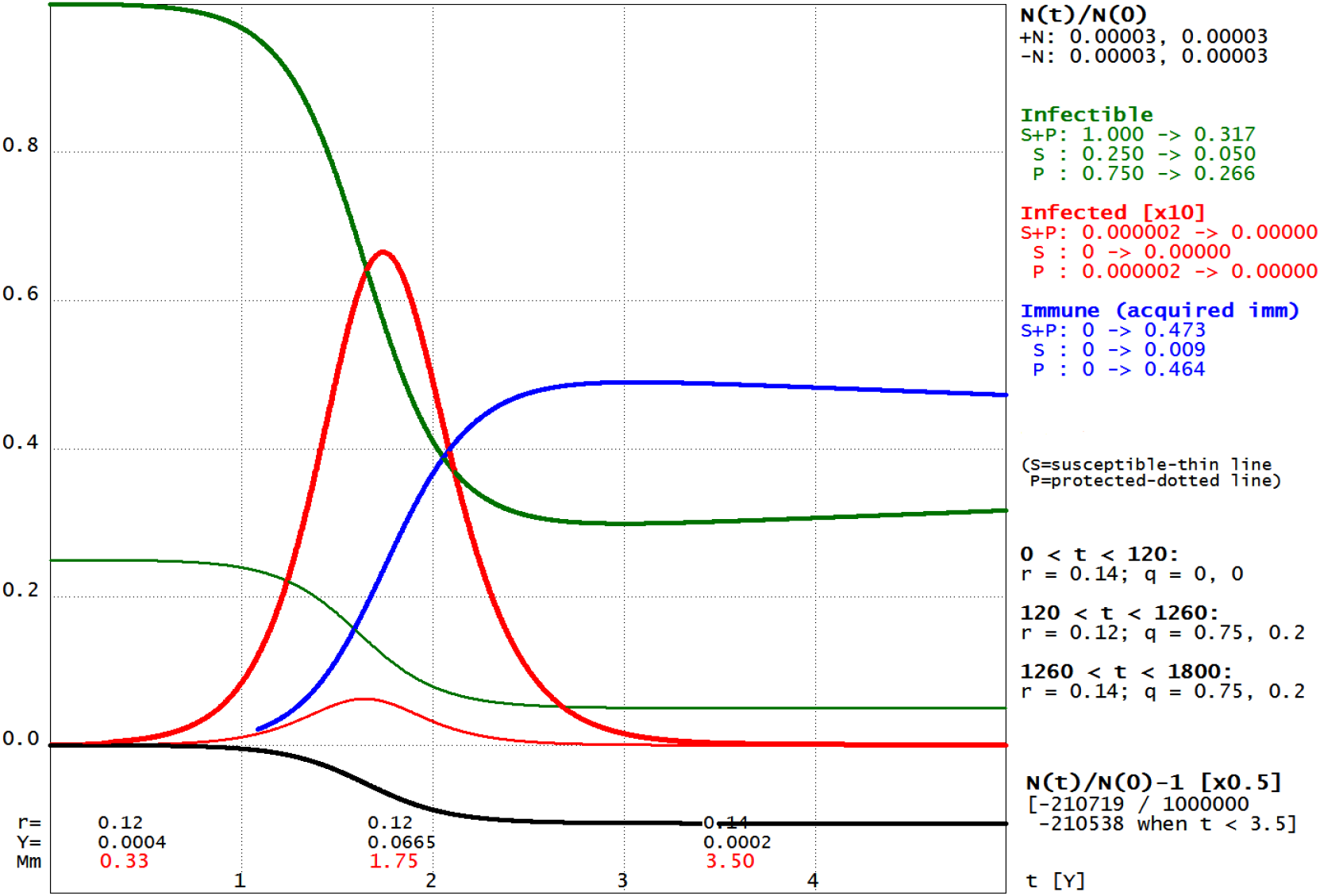
*x*_*S*_ (0) = 0.25, *x*_*P*_ (0) = 0.75. *r* (0) = 0.14; for *t* ≥ 120, *r* = 0.12, *q* = 0.75, 0.2.

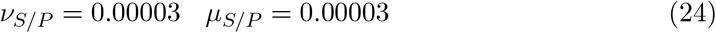

*per day* (= 0.0108 per year, Y; 1 Y = 360 D); *γ* is the same of Sections 5.1 and 5.2 while

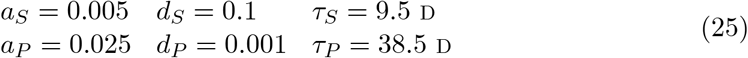

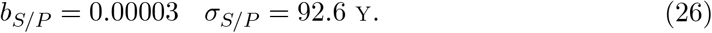

The dying probabilities (by the disease) of the susceptible and the protected are, respectively, equal to *d*_*S*_*τ*_*S*_ = 0.95 and to *d*_*P*_*τ*_*P*_ = 0.038.

Now *three quarters* of the population are (somewhat) protected and, when *t* = 0, there are 2 × 10^−6^*N*_0_ infected individuals (protected):

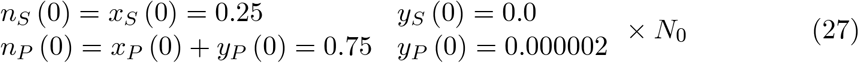

*z*_*S/P*_ (0) = 0. Moreover *r* = 0.14, when *t* ≤ 120 D and *r* = 0.12, *q* = 0.75, 0.2, when *t* > 120 D; *r* is reset to 0.14 when *t* = 3.5 Y. Hence:

Now *X* and *Z* will *oscillate*, as *Y*. But, in *this case, Y* rapidly reduces to negligible values: after 10 years (from the epidemic onset) *Y* ≅ 7 × 10^−1O^*N*_0_ and, at its first, absolute, minimum (after 15 years), *Y* ≅ 4 × 10^−11^*N*_0_; also if *N*_0_ ≅ 10^9^ the epidemic is, “for all practical purpose”, *definitely over* – if it is not reintroduced from the outside.

### 5.4 Endemic Phases

Diseases with rapid and efficient diffusion, but relatively *low mortality* rates, generally quickly reach an *endemic phase*: see **Fig. 4a**, which shows a *five years* continuation of **Fig. 2a**. Here it is assumed that:

**Fig. 4a.**
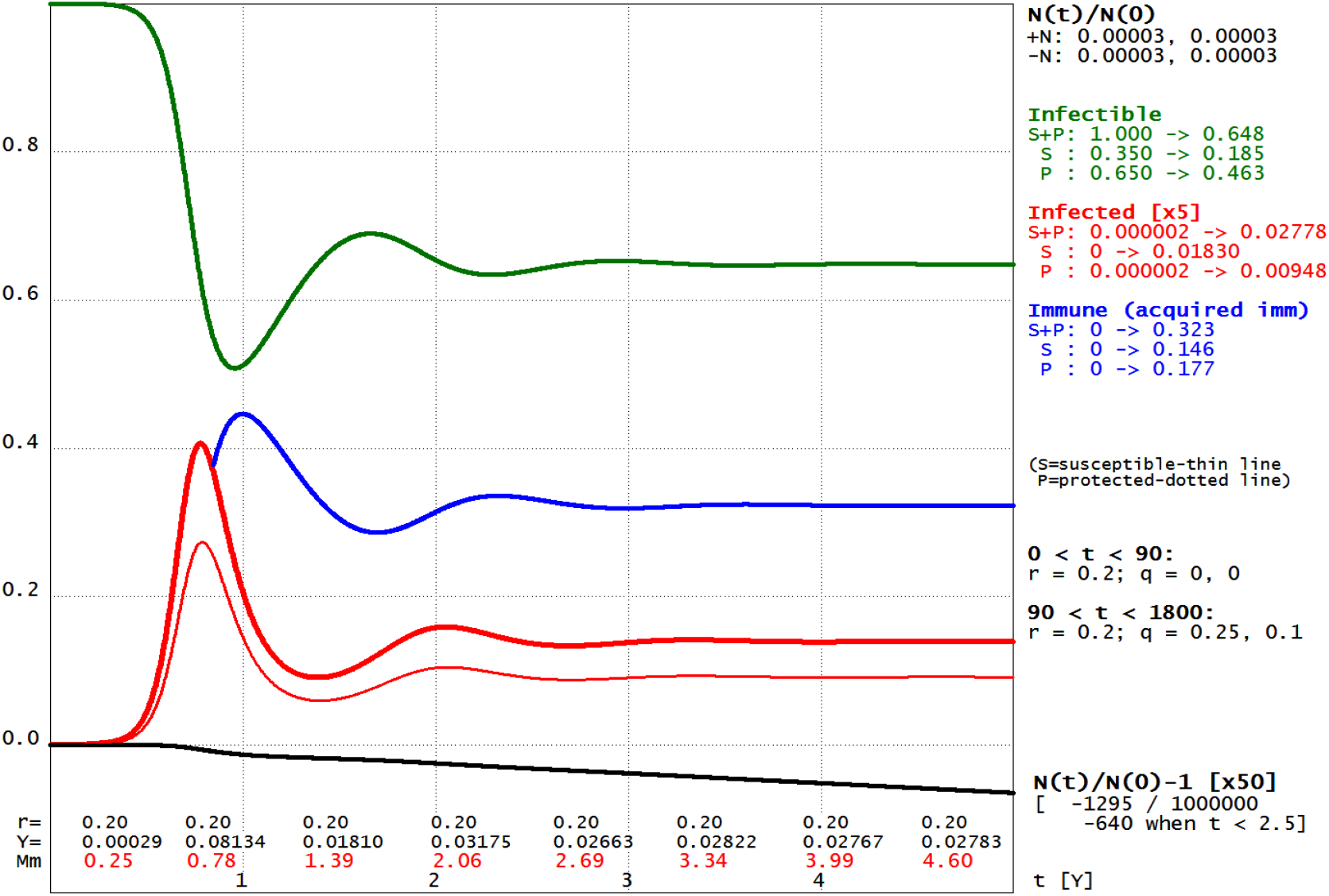
*x*_*S*_ (0) = 0.35, *x*_*P*_ (0) = 0.65. *r* is kept equal to 0.20, *q* = 0.25, 0.1.

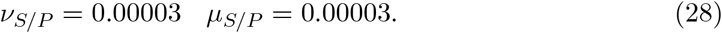

Things do not change significantly if, after an initial energetic intervention, the *r* and *q* are (inevitably) reset close to their initial values. See **Fig. 4b**, which shows a *five years* continuation of **Fig. 2c**: the endemic phase is only slightly *postponed*. And, when *t*_*M*_ = 5 Y, the values of *X, Y* and *Z* are, in both cases, almost the same:

**Fig. 4b.**
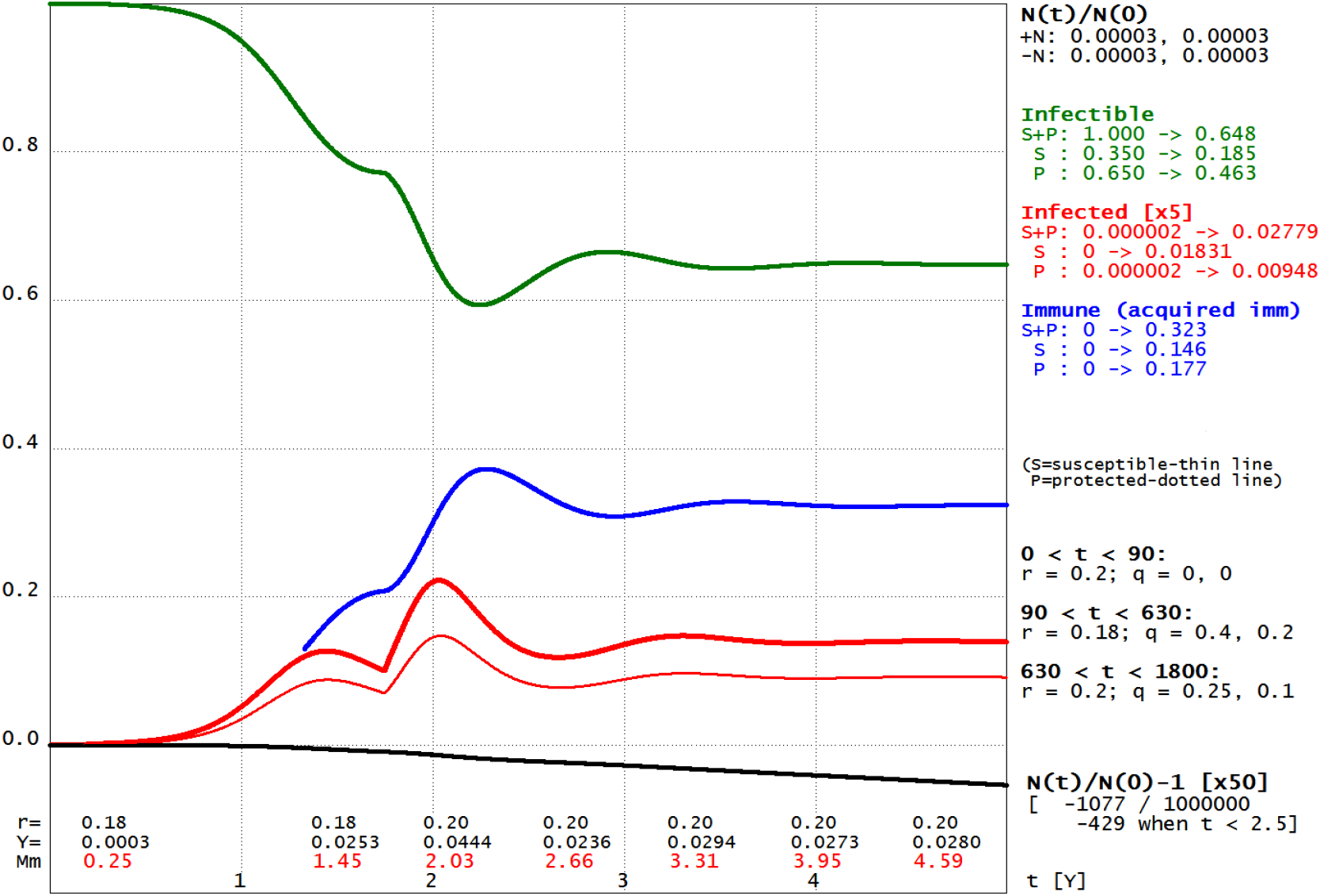
*x*_*S*_ (0) = 0.35, *x*_*P*_ (0) = 0.65. *r* (0) = 0.20; *r* will be restored when *t* = 630.

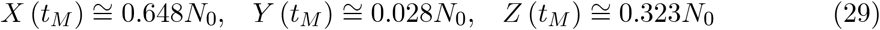

while *N* (*t*_*M*_) ≅ 0.999*N*_0_.

Generally the *deadly diseases* do not enter in the endemic phase, they die out (as in **Fig. 3a**), but it may happen that, for *very low values* of the contact rate *r*, the first minimum of the infected curve is still high enough, then the following minima of the curve will be *even higher* and the disease will move towards an endemic phase; see the characteristic **Fig. 4c**.

**Fig. 4c.**
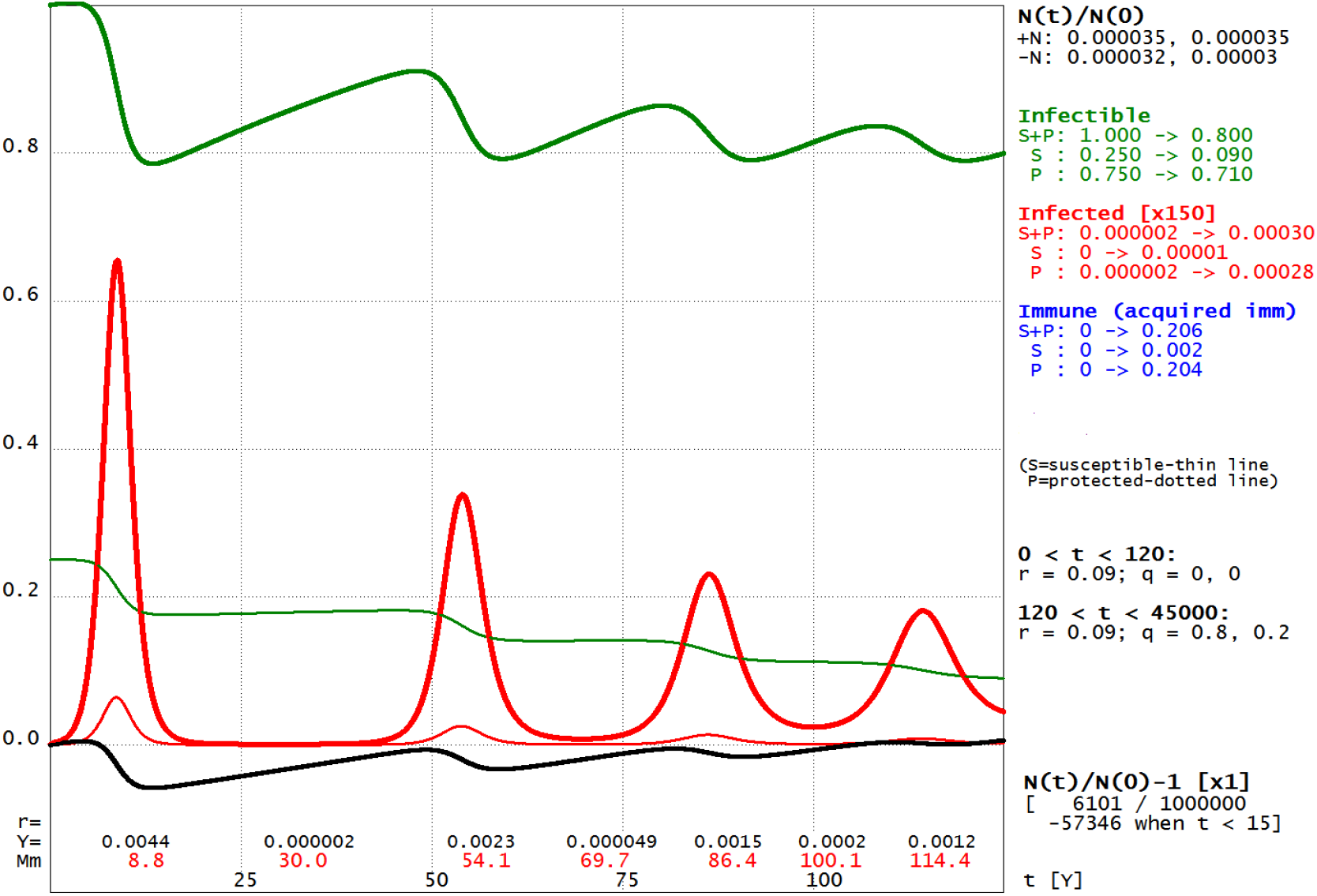
*x_S_* (0) = 0.25, *x*_*P*_ (0) = 0.75. *r* (0) = 0.09; for *t* ≥ 120, *r* = 0.09, *q* = 0.8, 0.2.

In this case

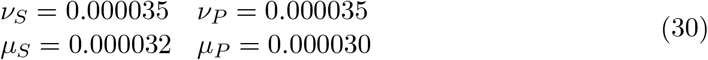

*per day*; they are chosen in such a way that, after 125 years, *N* (*t*) ≅ *N* (0). *γ* and the *a, b* and *d* are the same of Section 5.3, **Fig. 3a**.

Again *three quarters* of the population are protected and, when *t* = 0, there are 2 × 10^−6^*N*_0_ infected individuals, as in Section 5.3. But now it is always *r* = 0.09; *q*_*S*_ = 0.75, *q*_*P*_ = 0.2, for *t* > 120 D. Hence, after 125 years, *N* = 1.006*N*_0_ and

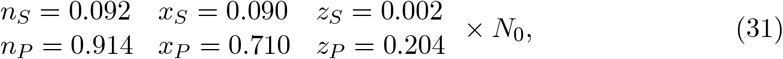

you can see that the susceptible are in the process of disappearing.

It may be convenient to define a sort of *measure of the endemicity* of the epidemic, at time t. For example:

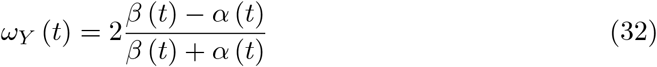

where *α* (*t*) = inf_*s*≥*t*_ *Y* (*s*) /*N* (*s*) and *β* (*t*) = sup_*s*≥*t*_ *Y* (*s*) /*N* (*s*). *ω*_*Y*_ (*t*) has to be “small”: in the case of **Fig. 4a**, *ω*_*Y*_ (*t*) = 0.02 when *t* = 3.3 Y while, for **Fig. 4b**, *ω*_*Y*_ (t) = 0.025 when *t* = 3.9 Y.

The “final” endemic phase will begin after almost 300 years, depending on the values of the *ν* and *μ*. Note that a casual observer, who has come into contact with the disease in its final endemic phase (and without other information), may be tempted to attribute the origin of the disease to “unknown causes” – or similarly.

### 5.5 The Tolerant

Now we shall assume that the protected individuals (*P*) are, in fact, *tolerant*: they *cohabit* (collaborate) with the agents of the disease and both the recovery and the disease death rate are = 0.

The infected tolerant are assumed to be slightly infectious:

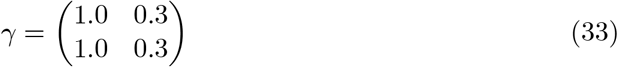

and

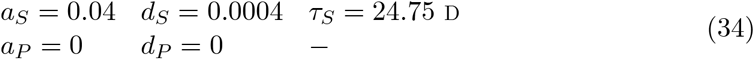

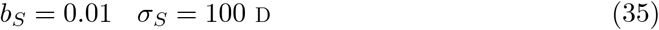

The probability (*d*_*S*_*τ*_*S*_) that a susceptible dies, after contracting the disease is = 0.01.

One quarter of the population is assumed to be tolerant; and, when *t* = 0, there are 10^−6^*N*_0_ infected (tolerant):

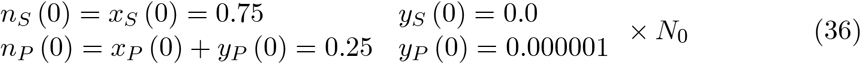

*z*_*S*_ (0) = 0 and, in this case (*a*_*P*_ = 0), *z*_*P*_ is always = 0. The contact rate *r* is now assumed to be *very low*, = 0.02; *q*_*S*_ = 0.50, *q*_*P*_ = 0.05, for *t* > 720 D.

The birth rates are quite high (but not enough to compensate for the disease death rate, for the susceptible):

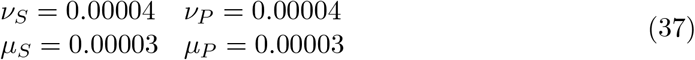

*per day* (0.0144 and 0.0108 per year); they are chosen in such a way that, after 300 years, *N* (*t*) ≅ *N* (0), see the **Fig. 5a**. Observe that *y*_*P*_ = *x*_*P*_ = 0.134*N*_0_ when *t* = 19.8 Y and *y*_*P*_ = *x*_*S*_ = 0.426*N*_0_ when *t* = 152.3 Y.

**Fig. 5a.**
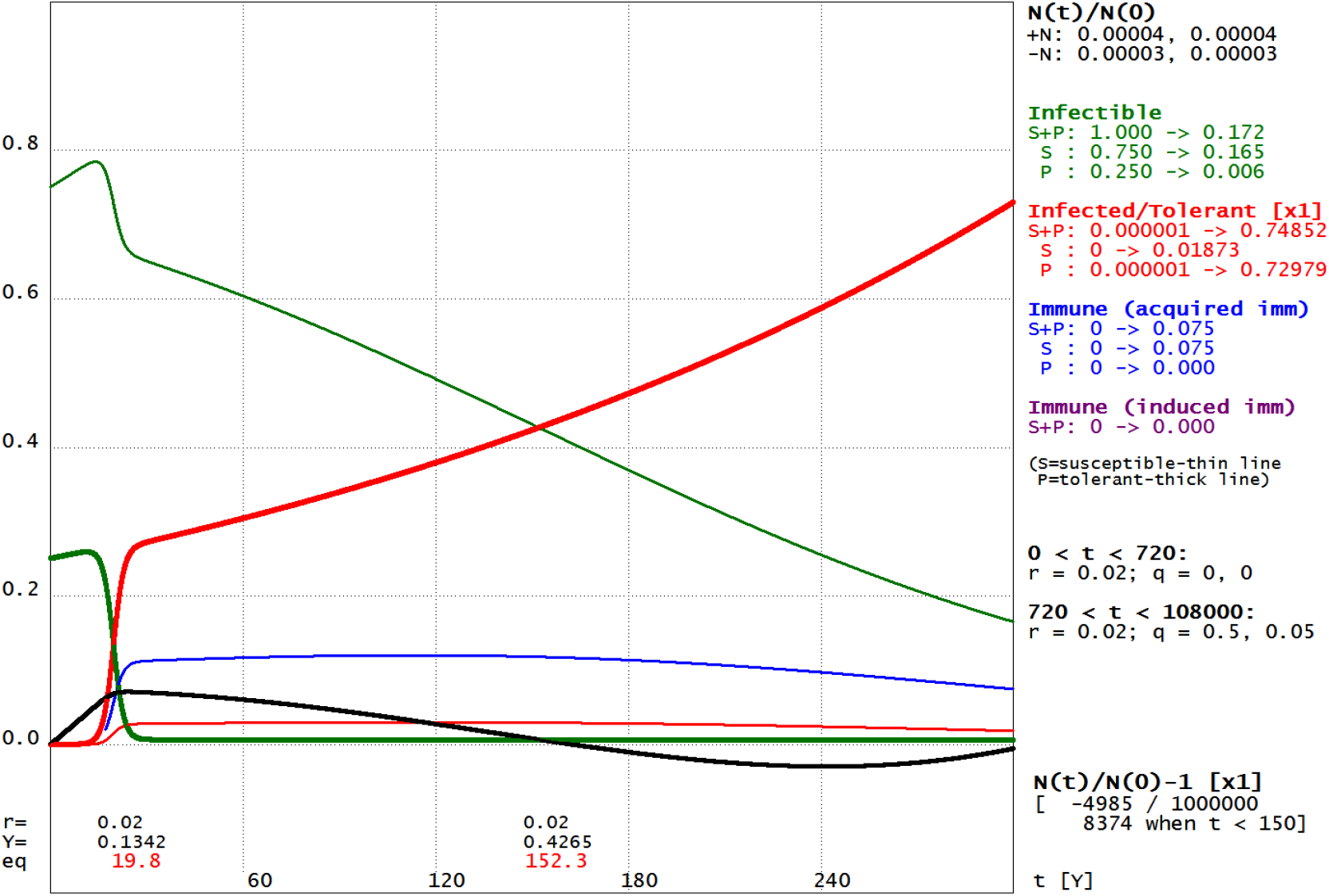
*x*_*S*_ (0) = 0.75, *x*_*P*_ (0) = 0.25. *r* is kept equal to 0.02, *q* = 0.5, 0.05.

Tolerance is assumed to be *congenital*, but the tolerant newborns are here assumed to be *uninfected*. On the other hand, if all of them are instead infected, the differential equations must be slightly modified:

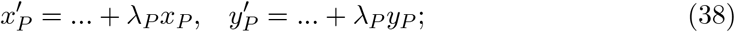

however things do not change significantly.

The presence of infectious tolerant obviously poses some social and ethical dilemma.

## Data Availability

No data

## Appendices

### A The infinite-dimensional case

The model can be extended in a obvious way assuming that *K* is a generic compact (then complete) metric space, that *dk* is a *positive measure* on *K* and that *t* ∈ *I* ↦ *x*_*t*_, *y*_*t*_ and *z*_*t*_ are continuously differentiable maps *I* → *E* = *L*^1^ (*K, dk*), the Banach space of (equivalence classes of) *dk*-summable functions *K* → ℝ. *n*_*t*_ = *x*_*t*_ + *y*_*t*_ + *z*_*t*_ and, if *H* is a measurable subset of *K*,

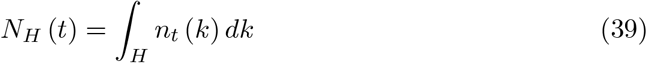

is the “number” of individuals characterized by *H. X*_*H*_ (*t*) = *∫*_*H*_ *x*_*t*_ (*k*) *dk, X* (*t*) = *X*_*K*_ (*t*) and likewise for *Y* and *Z*; *N* (*t*) = *X* (*t*) + *Y* (*t*) + *Z* (*t*).

Furthermore, for every *t* ∈ *I, r*_*t*_ is now a continuos map *K* x *K* ↦ ℝ, *r*_*t*_ (*h, k*) ≥ 0, *∫*_*K* × *K*_ *r*_*t*_ (*h, k*) *dhdk* > 0 and

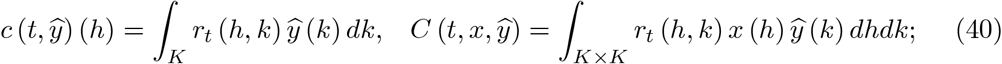

*r* and *ŷ* (as *a, b, d, q*) are defined as in the finite-dimensional case. We arrive at a system of differential equations in a infinite-dimensional Banach space; the theory of such systems does not differ significantly from the finite-dimensional theory; see, for example, L. Schwartz [9].

### B Differential inequalities

*I* is a real, open interval; *a* and *b* are *bounded* and *continuos* functions *I* ↦ ℝ, *a* ≤= *b*; *u* is a *continuous differentiable* function *I* ↦ ℝ, *u* ≥ 0. Then put 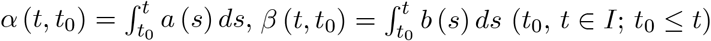 and assume that, on *I*,

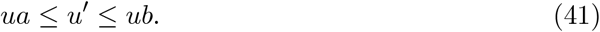

Now the functions 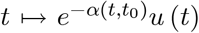 and 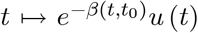 are *continuous differentiable* and their derivatives are respectively equal to 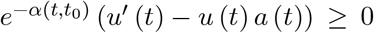 and to 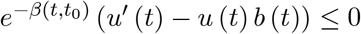 so, for *t* ≥ *t*_0_:

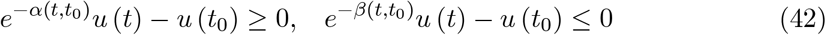

or

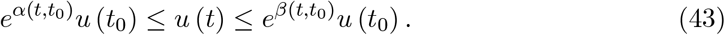

If *u* (*t*_0_) > 0, *u* (*t*) is > 0, for *t* ≥ *t*_0_; otherwise *u* (*t*) is always = 0.

Note that, if *u* ′ < *ub* or *ua* < *u* ′,

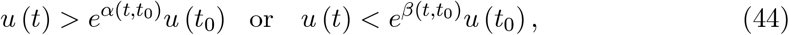

now only for *t* > *t*_0_.

In our case (Section 4) observe that (*h, k* ∈ *K, λ*_*k*_ = *ν*_*k*_ - *μ*_*k*_, *x*_*k*_ ≤ *n*_*k*_, *y*_*k*_ ≤ *n*_*k*_):

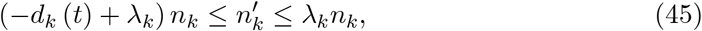

hence

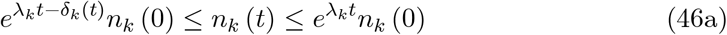

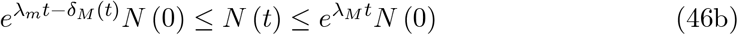

where 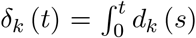 and, generally in the following, for *u* ∈ *E, u*_*m*_ = min_*k* ∈*K*_ *u*_*k*_, *u*_*M*_ = max_*k* ∈*K*_ *u*_*k*_. Consequently, if *n*_*k*_ (0) = 0, *n*_*k*_ (*t*) is always = 0; otherwise it is > 0 (as for *N*); obviously only the latter case is here of any interest. Note that, if *λ*_*k*_ = 0,

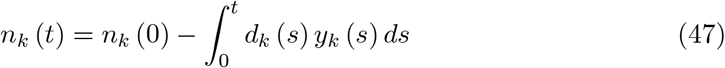

while, if *d*_*k*_ = 0, 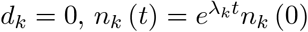, as expected.

#### The infectible

Observe now that, if *N* > 0, ∑_*k*_ *ŷ*_*k*_ = (*Y* − ∑_*k*_ *q*_*k*_*y*_*k*_) / (*N* − ∑_*k*_ *q*_*k*_*y*_*k*_) is ≤ *Y*/*N*, so

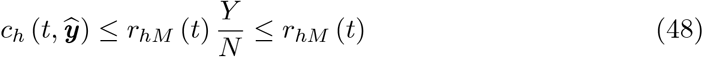

and

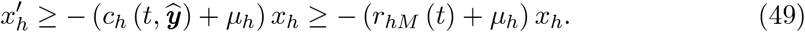

Hence

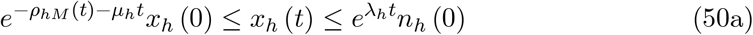

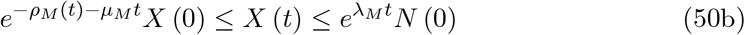

where 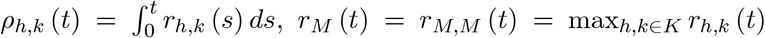. Consequently *x*_*h*_ (*t*) is always > 0 if *x*_*h*_ (0) > 0 (as for *X*); here it will be assumed that it is always so.

#### The infected

We need an upper bound for *C* (*t, x, ŷ*), depending on *Y*:

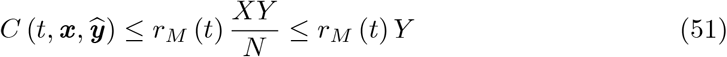

and (*ā*_*h*_ (*t*) := *a*_*h*_ (*t*) + *d*_*h*_ (*t*) + *μ*_*h*_)

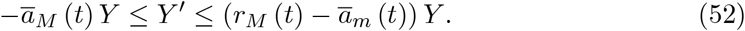

Hence

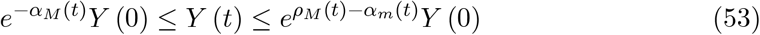

Where 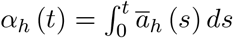. Consequently, if *Y* (0) = 0, *Y* (*t*) is always = 0; otherwise it is always > 0. Obviously only the latter case is here of interest; then some *y*_*h*_ (0) must be > 0.

Generally 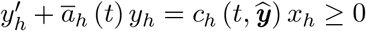, hence, if *y*_*h*_ (0) > 0,

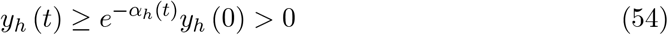

for *t* > 0. But if *y*_*h*_ (0) = 0 and *c*_*h*_ (*t, ŷ*) > 0 (for example, if all *r*_*h,k*_ (*t*), *k* ∈ *K*, are > 0),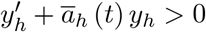 and

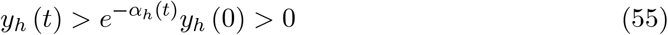

for *t* > 0. If instead *y*_*h*_ (0) = 0 and *c*_*h*_ (*t, ŷ*) = 0 (all *r*_*h,k*_ (*t*), *k* ∈ *K*, are = 0, say),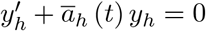 and

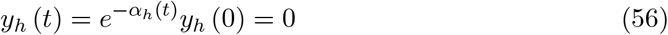

for *t* > 0. In this case also *z*_*h*_ (*t*) is always = 0 (see below, The immune) and 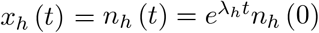.

Finally note that 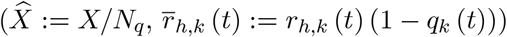:

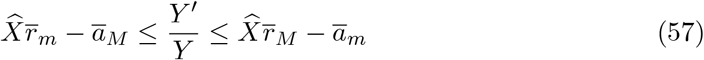

which shows that if, for some *t*,

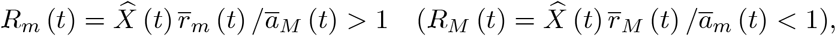

*Y* is strictly increasing (decreasing), at *t*. But for an *inhomogeneous* population (with respect to the disease) 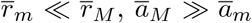 and, generally, *R*_*m*_ (*t*) − 1 and *R*_*M*_ (*t*) − 1 have a *different* sign.

#### The immune

Posing 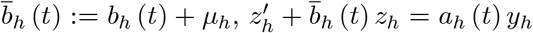. Now, if *z*_*h*_ =(0) and *a*_*h*_ (*t*) *y*_*h*_ (*t*) > 0,

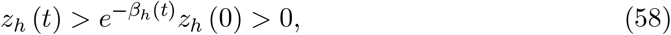

where 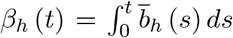. Consequently if, for some *h* ∈ *K, a*_*h*_ (*t*) *y*_*h*_ (*t*) > 0, *Z* (*t*) is > 0, for *t* > 0.

But if *z*_*k*_ (0) = 0 and *a*_*h*_ = 0 (a deadly disease, if *d*_*h*_ > 0) or *y*_*h*_ = 0 (fully protected individuals)

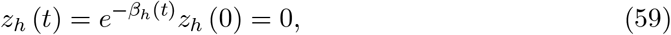

as expected.

